# Simulation and predictions of a new COVID-19 pandemic wave in Ukraine with the use of generalized SIR model

**DOI:** 10.1101/2021.10.13.21264949

**Authors:** Igor Nesteruk

**Affiliations:** Institute of Hydromechanics, National Academy of Sciences of Ukraine, Kyiv, Ukraine; Igor Sikorsky Kyiv Polytechnic Institute, Kyiv, Ukraine

**Keywords:** COVID-19 pandemic, epidemic dynamics in Ukraine, mathematical modeling of infection diseases, SIR model, parameter identification, statistical methods

## Abstract

A new wave of the COVID-19 pandemic in Ukraine, which began in the summer of 2021, was characterized by almost exponential growth of smoothed daily numbers of new cases. This is a matter of great concern and the need to immediately predict the dynamics of the epidemic in order to assess the possible maximum values of new cases, the risk of infection and the number of deaths. The generalized SIR-model and corresponding parameter identification procedure was used to simulate and predict the dynamics of this epidemic wave. The new COVID-19 epidemic wave in Ukraine will begin to subside in mid-October 2021, but its duration will be quite long. Unfortunately, new cases may appear by the summer of 2022.

## Introduction

The COVID-19 pandemic dynamics in Ukraine was discussed in [1-14]. To predict the first wave of the pandemic, the classical SIR model [15-17] and the statistics-based method of its parameter identification [18] were used. To simulate new epidemic waves, a numerical method of their detection [3, 19], a generalized SIR-model [20] and a corresponding parameter identification procedure [21] were developed. In particular, eleven epidemic waves in Ukraine were calculated [4, 7-10].

The calculations of the 11-th pandemic wave (based on the accumulated numbers cases reported by Ukrainian national statistics [22, 23] in the period May 23 – June 5, 2021) predicted the end of this wave on August 25, 2021 with the number of cases 2,226,797 (see [10]). As of August 25, 2021 the real number of cases accumulated in Ukraine was 2,278,171 (see Table 1). It means that the predicted saturation level was exceeded only 2.26% (after 81 days of observation). The obtained high accuracy of the method allows us to hope for a fairly accurate forecast for the next 12th epidemic wave in Ukraine, to which this study is devoted.

**Table 1.** Cumulative numbers of laboratory-confirmed Covid-19 cases in Ukraine *V*_*j*_ in the summer and autumn of 2021 according to the national statistics, [22, 23].

| Day in corresponding month of 2021 | Number of cases in May, $V_j$ | Number of cases in June, $V_j$ | Number of cases in July, $V_j$ | Number of cases in August, $V_j$ | Number of cases in September, $V_j$ | Number of cases in October, $V_j$ |
| --- | --- | --- | --- | --- | --- | --- |
| 1 | 2083180 | 2206836 | 2236497 | 2253534 | 2290848 | 2447222 |
| 2 | 2085938 | 2209417 | 2237202 | 2254361 | 2293541 | 2455189 |
| 3 | 2088410 | 2211683 | 2237579 | 2255345 | 2296155 | 2460010 |
| 4 | 2090986 | 2213580 | 2237823 | 2256397 | 2297534 | 2469856 |
| 5 | 2097024 | 2214517 | 2238364 | 2257478 | 2298307 | 2482518 |
| 6 | 2105428 | 2215052 | 2238974 | 2258532 | 2300504 | 2497643 |
| 7 | 2114138 | 2216654 | 2239591 | 2259151 | 2303276 | 2514005 |
| 8 | 2119510 | 2218039 | 2240246 | 2259451 | 2306939 | 2529913 |
| 9 | 2122327 | 2219824 | 2240753 | 2260232 | 2310554 | 2541257 |
| 10 | 2124535 | 2221427 | 2241043 | 2261354 | 2314423 | 2550089 |
| 11 | 2129073 | 2222701 | 2241217 | 2262601 | 2316619 | 2562085 |
| 12 | 2135886 | 2223558 | 2241698 | 2263864 | 2317824 | 2578394 |
| 13 | 2143448 | 2223978 | 2242245 | 2265217 | 2321156 | - |
| 14 | 2150244 | 2224992 | 2242868 | 2265912 | 2325796 | - |
| 15 | 2153864 | 2226037 | 2243605 | 2266329 | 2331540 | - |
| 16 | 2156000 | 2227225 | 2244196 | 2267219 | 2338164 | - |
| 17 | 2160095 | 2228192 | 2244495 | 2268666 | 2344398 | - |
| 18 | 2165233 | 2229044 | 2244677 | 2270226 | 2348381 | - |
| 19 | 2170398 | 2229523 | 2245275 | 2271826 | 2350646 | - |
| 20 | 2175382 | 2229846 | 2245930 | 2273558 | 2355805 | - |
| 21 | 2179988 | 2230142 | 2246656 | 2274561 | 2362559 | - |
| 22 | 2182521 | 2230977 | 2247419 | 2275171 | 2370425 | - |
| 23 | 2183855 | 2231914 | 2248164 | 2275863 | 2379483 | - |
| 24 | 2186463 | 2232790 | 2248450 | 2276590 | 2387750 | - |
| 25 | 2189858 | 2233546 | 2248663 | 2278171 | 2392397 | - |
| 26 | 2193367 | 2233996 | 2249344 | 2280203 | 2395404 | - |
| 27 | 2196673 | 2234281 | 2250061 | 2282285 | 2401956 | - |
| 28 | 2199769 | 2234463 | 2250907 | 2284191 | 2411622 | - |
| 29 | 2201472 | 2235096 | 2251869 | 2284940 | 2423379 | - |
| 30 | 2202494 | 2235801 | 2252785 | 2286296 | 2435413 | - |
| 31 | 2204631 | - | 2253269 | 2288371 | - | - |

### Data

We will use the data set regarding the accumulated numbers of laboratory-confirmed COVID-19 cases in Ukraine from national sources [22, 23]. The corresponding numbers *V*_*j*_ and moments of time *t*_*j*_ (measured in days) are shown in Table 1. The values for the period *T*_*c11*_: May 23 – June 5, 2021 have been used in [10] for SIR simulations of the tenth epidemic wave in Ukraine. Here we use the dataset, corresponding to the period *T*_*c12*_: September 29 – October 12, 2021 to simulate the 12-th wave. Other *V*_*j*_ and *t*_*j*_ values will be used to control the accuracy of predictions.

To estimate the mortality rate (ratio of accumulated number of cases *V*_*j*_ to the accumulated number of deaths *D*_*j*_), let us take figures *D*_*j*_ corresponding different days: 52,286 (June 26, 2021); 52,665 (July 13, 2021); 52,981 (August 2, 2021); 53,789 (August 30, 2021); 54,550 (September 15, 2021); 57,526 (October 5, 2021); 59,052 (October 11, 2021), [22, 23]. Taking corresponding *V*_*j*_ values from Table 1, we can calculate the mortality rates *m*_*i*_ =*D*_*j*_ *1000/*V*_*j*_ (per thousand of cases) for the listed days: 23.40; 23.49; 23.50; 23.52; 23.40; 23.17; 23.05. Thus the mortality rate is rather stable (its variation during June to October 2021 is only 0.47). We will use the average value *m*= 23.36 to predict the number of deaths in Ukraine during the new 12-th pandemic wave.

### Generalized SIR model and data smoothing procedure

The generalized SIR-model relates the number of susceptible *S*, infectious *I* and removed persons *R* for a particular epidemic wave *i* [8, 20]. The exact solution of the set of non-linear differential equations uses the function

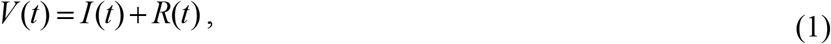

corresponding to the number of victims or the cumulative confirmed number of cases [8, 20]. Its derivative:

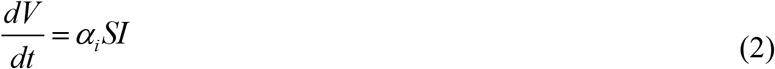

yields the estimation of the average daily number of new cases. When the registered number of victims *V*_*j*_ is a random realization of its theoretical dependence (1), the exact solution presented in [8, 20] depends on five parameters (*α*_*i*_ is one of them). The details of the optimization procedure for their identification can be found in [21].

Since daily numbers of new cases are random and characterized by some weekly periodicity, we will use the smoothed daily number of accumulated cases:

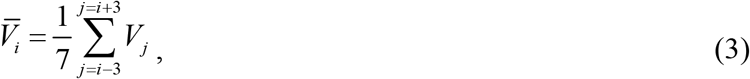

and its numerical derivative:

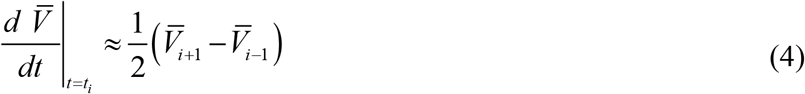

to estimate the smoothed number of new daily cases [3, 4, 9, 19].

## Results and discussion

The optimal values of the general SIR model and other characteristics of the 12-th pandemic wave in Ukraine are calculated and listed in Table 2 (middle column). The same characteristics of the 11-th wave calculated in [10] are presented in the last column of Table 2 for comparison. The corresponding SIR curves are shown in Figure by blue and black lines. It can be seen that the optimal values of SIR parameters are very different for 11-th and 12-th pandemic waves. Close values were obtained only for the average times of spreading the infection 1/ *ρ*_11_ =4.1 days (in comparison with 1/ *ρ*_12_ =3.3 days for the twelfth wave). The assessment of the 12-th epidemic wave duration corresponding the moment when the number of infectious persons becomes less that unit) is very pessimistic (June 16, 2022). A similar long epidemic wave was also predicted for India [24].

**Figure.**
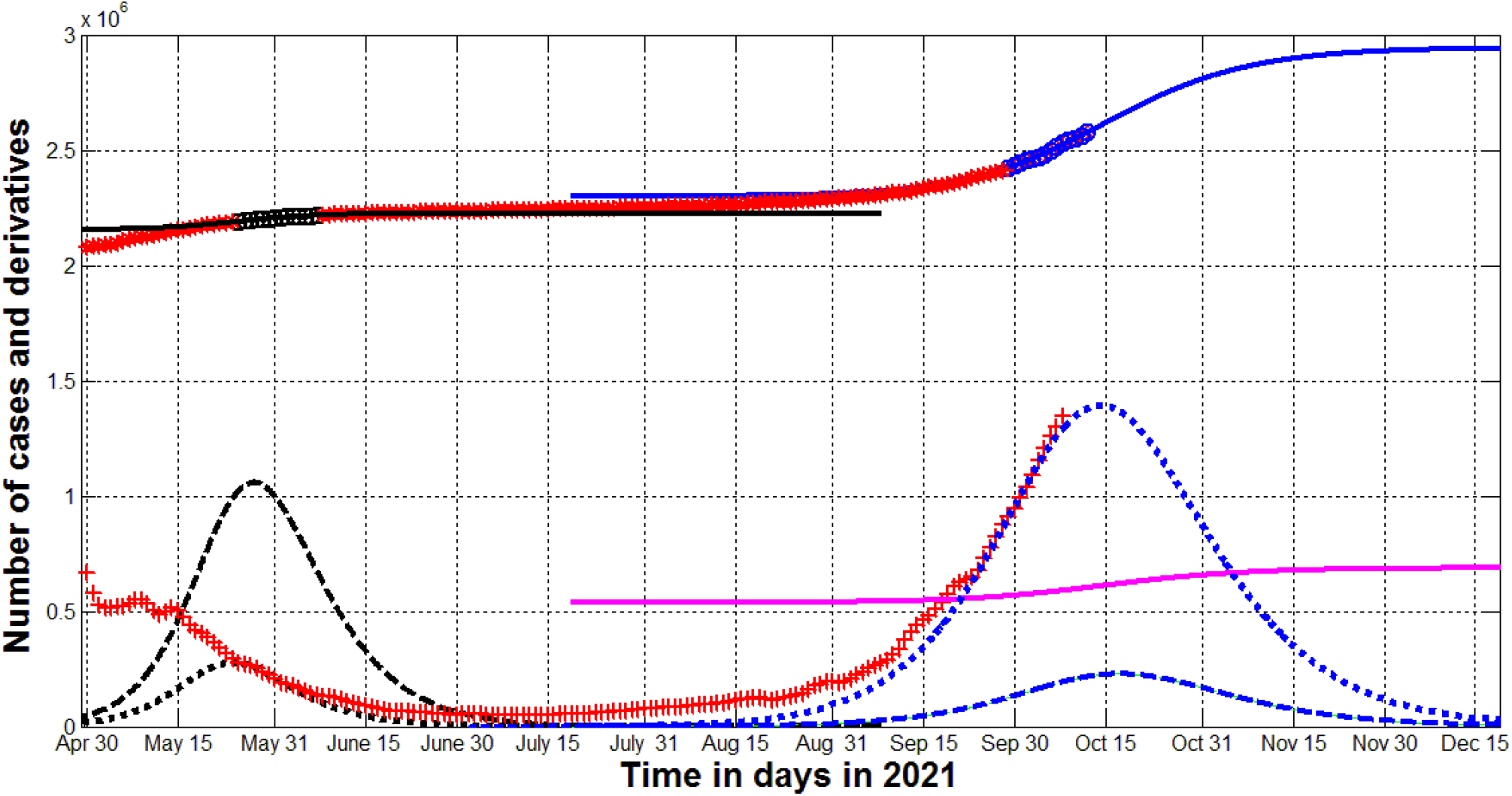
11-th and 12-th waves of the COVID-19 pandemic in Ukraine in the summer and autumn of 2021. The results of SIR simulations of the eleventh [10] and twelfth waves are shown by black and blue lines respectively. Numbers of victims *V(t)=I(t)+R(t)* – solid lines; numbers of infected and spreading *I(t)* multiplied by 5 – dashed; derivatives *dV/dt* (eq. (2)) multiplied by 100 – dotted. The magenta line represents the estimation of the accumulated number of deaths during the 12-th epidemic wave multiplied by 10. Black and blue “circles” correspond to the accumulated numbers of cases registered during the periods of time taken for SIR simulations (May 23 – June 5, 2021) and September 29 – October 12, 2021, respectively. “Stars” corresponds to *V*_*j*_ values beyond these time periods. The red “crosses” show the first derivative (4) multiplied by 100.

**Table 2.** Optimal values of parameters and other characteristics of the eleventh and twelfth COVID-19 pandemic wave in Ukraine.

| Characteristics | 12<sup>th</sup> epidemic wave,<br>$i=12$ | 11<sup>th</sup> epidemic wave,<br>$i=11$,<br>[10] |
| --- | --- | --- |
| Time period taken<br>for calculations $T_{ci}$ | September 29 – October 12,<br>2021 | May 23 – June 5, 2021 |
| $I_i$ | 25,261.4089164122 | 10,190.8327995721 |
| $R_i$ | 2,399,050.73394073 | 2,180,167.16720043 |
| $N_i$ | 3,790,400 | 2,258,464 |
| $\nu_i$ | 1,137,541.61656928 | 60,891.3982283695 |
| $\alpha_i$ | 2,63670321714285e-07 | 3.98540619731056e-06 |
| $\rho_i$ | 0.299935964004210 | 0.242676955862249 |
| $1/\rho_i$ | 3.33404499630449 | 4.12070440082342 |
| $r_i$ | 0.997536473683354 | 0.996838194153353 |
| $S_{i\infty}$ | 845,264 | 31,667 |
| $V_{i\infty}$ | 2,945,136 | 2,226,797 |
| Final day of the<br>epidemic wave | June 16, 2022 | August 25, 2021 |

The saturation level (final size) of the 12-th wave can approach 3 millions. As of the end of 2021 the accumulated numbers of laboratory-confirmed cases and deaths can exceed 2,943,700 and 68,764, respectively (see blue and magenta solid lines in Figure). The numbers of infectious persons and average daily new cases (see blue dashed and dotted lines) will stop to increase around 17 and 14 October 2021, respectively with the maximal values of 45,545 and 13,930, respectively (see blue dashed and dotted lines). As of the end of 2021 these values are expected to be around 505 and 113, respectively (see blue dashed and dotted lines).

To illustrate the accuracy of simulations the accumulated number of cases (red “stars”) and averaged daily numbers of new cases (eq. (4), red “crosses”) are also presented in Figure. Comparisons with corresponding solid and dotted lines show that the theoretical curves are in good agreement with the observations of the pandemic dynamics (markers). To estimate the accumulated number of deaths during the 12-th wave, we have multiplied the theoretical *V(t)=I(t)+R(t)* values by the coefficient *m*= 23.36 calculated before. The magenta line represents expected accumulated numbers of deaths multiplied by 10.

## Conclusions

The new COVID-19 epidemic wave in Ukraine will begin to subside in mid-October 2021, but its duration will be quite long. Unfortunately, new cases may appear by the summer of 2022.

## Data Availability

data is listed in the text

## Acknowledgements

The author is grateful to Oleksii Rodionov for his help in collecting and processing data.

## Notes

### Competing Interest Statement

The authors have declared no competing interest.

### Funding Statement

no funding

## References

1. Nesteruk I., Kydybyn I., Demelmair G. GLOBAL STABILIZATION TRENDS OF COVID-19 PANDEMIC. KPI Science News. 2020. No. 2. pp. 55–62. DOI: 10.20535/kpi-sn.2020.2.205124

2. Nesteruk I. Simulations and predictions of COVID-19 pandemic with the use of SIR model. Innov Biosyst Bioeng. 2020. vol. 4. no. 2. 110–121. doi: 10.20535/ibb.2020.4.2.204274. http://ibb.kpi.ua/article/view/204274

3. Nesteruk I. Coronasummer in Ukraine and Austria. [Preprint.] ResearchGate. 2020 June. DOI: 10.13140/RG.2.2.32738.56002

4. Nesteruk I. COVID-19 pandemic dynamics. Springer Nature. 2021. DOI: 10.1007/978-981-33-6416-5. https://link.springer.com/book/10.1007/978-981-33-6416-5

5. Pardhan S,. Drydakis N. Associating the Change in New COVID-19 Cases to GDP per Capita in 38 European Countries in the First Wave of the Pandemic. Front Public Health. 2021 Jan 20;8:582140. doi: 10.3389/fpubh.2020.582140.

6. Chintala S., Dutta R., Tadmor D. COVID-19 spatiotemporal research with workflow-based data analysis. Infect Genet Evol. 2021 Mar;88:104701. doi: 10.1016/j.meegid.2020.104701.

7. Nesteruk I.. Benlagha N. PREDICTIONS OF COVID-19 PANDEMIC DYNAMICS IN UKRAINE AND QATAR BASED ON GENERALIZED SIR MODEL. Innov Biosyst Bioeng. 2021. vol. 5. no. 1. p. 37–46. DOI: 10.20535/ibb.2021.5.2.230487 http://ibb.kpi.ua/article/view/230487

8. Nesteruk I. Visible and real sizes of new COVID-19 pandemic waves in Ukraine Innov Biosyst Bioeng. 2021. vol. 5. no. 2. pp. 85–96. DOI: 10.20535/ibb.2021.5.2.230487 http://ibb.kpi.ua/article/view/230487

9. Nesteruk I. Detections and SIR simulations of the COVID-19 pandemic waves in Ukraine. Comput. Math. Biophys. 2021;9:46–65. https://doi.org/10.1515/cmb-2020-0117

10. Nesteruk I. Will a natural collective immunity of Ukrainians restrain new COVID-19 waves? MedRxiv, July 2021. DOI: 10.1101/2021.07.20.21260840

11. Nesteruk I, Rodionov O. The impact of demographic factors on the accumulated number of COVID-19 cases per capita in Europe and the regions of Ukraine in the summer of 2021. medRxiv, July 2021. DOI: 10.1101/2021.07.04.21259980

12. Nesteruk I, Rodionov O, Nikitin AV. The impact of seasonal factors on the COVID-19 pandemic waves. medRxiv, August 2021. DOI:10.1101/2021.08.06.21261665

13. Nesteruk I. Comparison of the COVID-19 pandemics dynamics in Ukraine and Israel in the summer of 2021. Research Gate, September 2021. https://www.researchgate.net/publication/354497030_Comparison_of_the_COVID-19_pandemics_dynamics_in_Ukraine_and_Israel_in_the_summer_of_2021

14. Nesteruk I. Effects of testing and vaccination levels on the dynamics of the COVID-19 pandemic and the prospects for its termination. MedRxiv, September 2021. DOI: https://doi.org/10.1101/2021.09.20.21263823

15. Kermack WO, McKendrick AG. A Contribution to the mathematical theory of epidemics. J Royal Stat Soc Ser A. 1927;115:700–21.

16. Murray JD. Mathematical Biology I/II. New York: Springer; 2002.

17. Langemann D, Nesteruk I, Prestin J. Comparison of mathematical models for the dynamics of the Chernivtsi children disease. Mathematics in Computers and Simulation. 2016;123:68–79. DOI: 10.1016/j.matcom.2016.01.003

18. Nesteruk I. Statistics based models for the dynamics of Chernivtsi children disease. Naukovi Visti NTUU KPI. 2017;5:26–34. DOI: 10.20535/1810-0546.2017.5.108577

19. Nesteruk I. Identification of the New Waves of the COVID-19 Pandemic. In book: COVID-19 Pandemic Dynamics, Springer Nature, 2021. DOI: 10.1007/978-981-33-6416-5_8. https://link.springer.com/chapter/10.1007/978-981-33-6416-5_8

20. Nesteruk I. General SIR Model and Its Exact Solution. In book: COVID-19 Pandemic Dynamics, Springer Nature, 2021. DOI: 10.1007/978-981-33-6416-5_9. https://link.springer.com/content/pdf/10.1007%2F978-981-33-6416-5_9.pdf

21. Nesteruk I. Procedures of Parameter Identification for the Waves of Epidemics. In book: COVID-19 Pandemic Dynamics, Springer Nature, 2021. DOI: 10.1007/978-981-33-6416-5_10. https://link.springer.com/chapter/10.1007%2F978-981-33-6416-5_10

22. Coronavirus in Ukraine - Statistics - Map of infections, graphs [Internet]. Index.minfin.com.ua. 2021. https://index.minfin.com.ua/ua/reference/coronavirus/ukraine/

23. Cabinet of Ministers of Ukraine – Home [Internet]. https://www.kmu.gov.ua/

24. Nesteruk I. The COVID-19 pandemic storm in India. [Preprint] medRxiv Posted May 08, 2021. doi: https://doi.org/10.1101/2021.05.06.21256523

